# Deep Learning in Magnetic Resonance Enterography for Crohn’s Disease Assessment: A Systematic Review

**DOI:** 10.1101/2023.12.27.23300507

**Authors:** Ofir Brem, David Elisha, Eli Konen, Michal Amitai, Eyal Klang

## Abstract

Crohn’s disease (CD) poses significant morbidity, underscoring the need for effective, non-invasive inflammatory assessment using magnetic resonance enterography (MRE).

This literature review evaluates recent publications on deep learning’s role in enhancing MRE segmentation, image quality, and visualization of inflammatory activity related to CD.

We searched MEDLINE/PUBMED for studies that reported the use of deep learning algorithms for assessment of CD activity. The study was conducted according to the PRISMA guidelines. The risk of bias was evaluated using the QUADAS_J2 tool.

Five eligible studies, encompassing 468 subjects, were identified.

Our study suggests that diverse deep learning applications, including image quality enhancement, bowel segmentation, and motility measurement are useful and promising for CD assessment. However, most of the studies are preliminary, retrospective studies, and have a high risk of bias in at least one category.

Future research is needed to assess how automated deep learning can impact patient care, especially when considering the increasing integration of these models into hospital systems.

## Introduction

Crohn’s disease (CD) is associated with substantial morbidity^1,2^. The management of inflammation is crucial in preventing disease complications - emphasizing the importance of effective assessment of inflammation^3^.

Colonoscopy stands as the gold standard for CD diagnosis. However, it is invasive, and the evaluation of the small bowel remains inadequate^4^. Magnetic Resonance Enterography (MRE) is a non-invasive technique that is effective for assessment of CD activity^4–6^ ^7,8^. MRE’s noninvasive nature also offers potential in evaluating treatment response, or identifying therapeutic inefficiency. This can prompt early detection and timely adjustments in therapy to maintain clinical remission^9–11^. Nevertheless, diagnosing Crohn’s disease using MRE is time-intensive and demands high expertise.

In recent years, AI, especially convolutional neural networks (CNN), have notably impacted computer vision. CNNs, a type of deep learning (**Figure 1**), excel in pattern recognition and are affecting the way in which medical images can be analyzed^12,13^. This technology offers an innovative approach to diagnosing and monitoring CD activity^14–17^.

**Figure 1.**
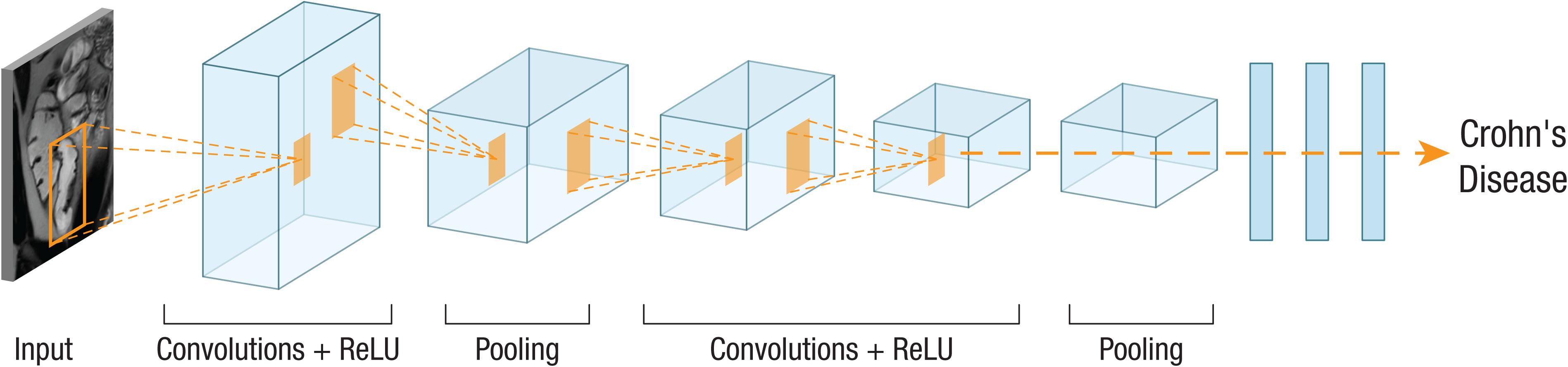
CNN specialize in image processing, utilizing small filters per layer to identify recurring patterns. Their hierarchical structure enables shallow layers to detect low-level patterns and deeper layers to grasp high-level image comprehension.

We reviewed the literature to evaluate articles focused on the use of deep learning to improve MRE analysis in Crohn’s disease.

## Methods

This review adhered to the guidelines outlined in the Preferred Reporting Items for Systematic Reviews and Meta-Analyses (PRISMA). The PRISMA checklist for systematic reviews can be found in **Supplementary Material 1.**

### Search strategy

We conducted an extensive literature search on October 1, 2023, using the PubMed/MEDLINE database to identify studies investigating the use of deep learning for detecting CD in MRI/MRE Our search terms included “MRI or MRE,” and “Crohn’s disease,” and “deep learning,” or related terms such as “convolutional neural networks,” “machine learning,” and “artificial intelligence.”

Detailed information about the complete search strategies can be found in **Supplementary Material 2.** We also conducted a manual search of the references in the studies we included.

Inclusion criteria encompassed studies that (1) assessed the effectiveness of a deep learning model in detecting CD on MRI/MRE, (2) were published in the English language, (3) were peer-reviewed original publications, and (4) included an outcome measure.

Exclusions were applied to articles not related to computer vision, non-deep learning articles, non-original articles, and abstracts.

This study is registered with PROSPERO under the registration number CRD42023484725.

### Study selection

Two authors (OB and EK) autonomously assessed the titles and abstracts to ascertain if the studies satisfied the inclusion criteria. When the title met the inclusion criteria or if any uncertainty arose, a thorough examination of the full-text article was conducted. If a relevant title appeared in the references section of one of the included studies, it was also screened for inclusion. In cases of disagreements, a third reviewer (DE) was consulted for resolution.

### Data extraction

Utilizing a uniform data extraction template in Microsoft Excel, the two reviewers (OB and EK) separately gathered information. The data encompassed details such as publication year, study design and location, patient count, ethical considerations, inclusion and exclusion criteria, study population description, deep learning technique, utilization of an online database, database size, incorporation of an independent test dataset, performance of cross-validation, assessment metrics employed, and the main findings.

### Quality assessment and risk of bias

We evaluated the quality of the studies using the modified Quality Assessment of Diagnostic Accuracy Studies (QUADAS-2) criteria^18^.

## Results

### Study selection and characteristics

The initial literature search resulted in 16 articles. Five studies met our inclusion criteria (**Supplementary Figure 1**). Studies were published between 2019 and 2023. A total of 468 subjects were analyzed. **Table 1** summarizes the characteristics of the included studies. **Figure 2** summarizes the clinical application of each study included.

**Figure 2.**
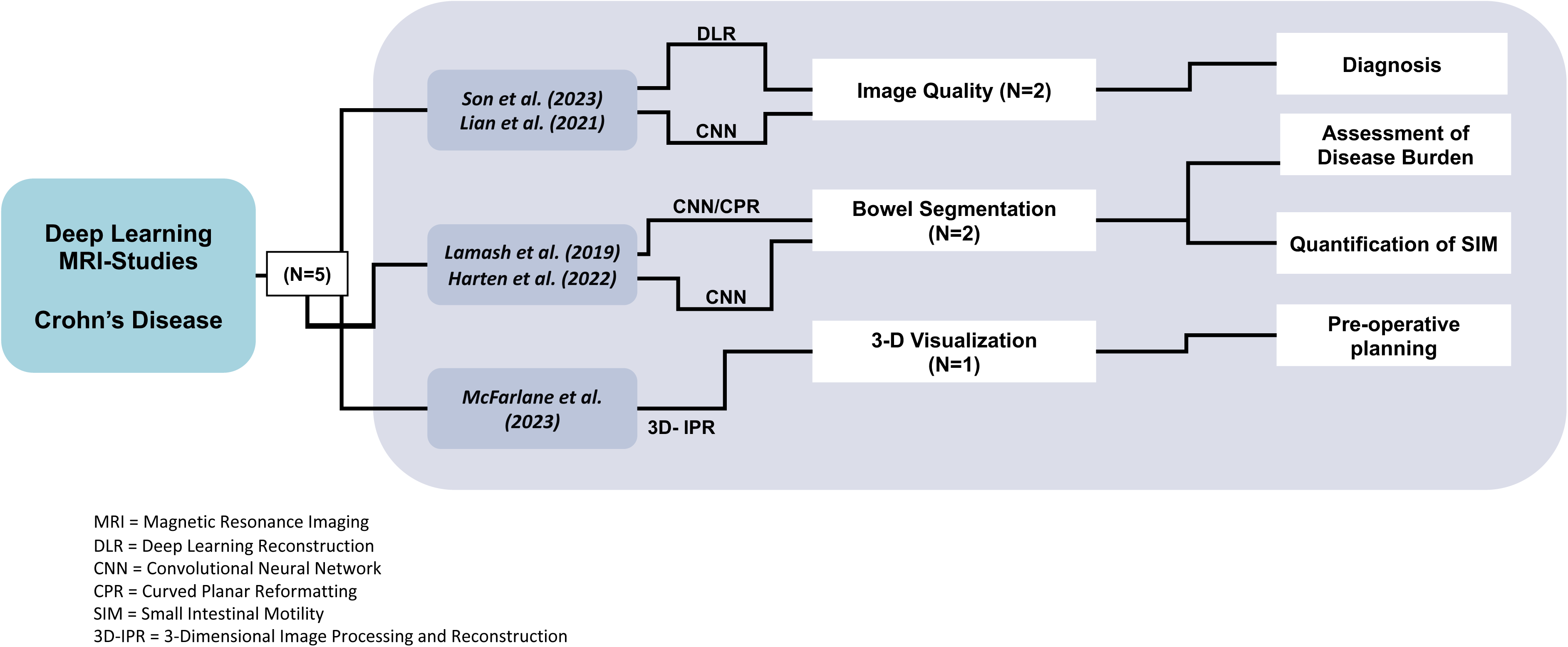
Graphical depiction categorizing deep learning studies according to their clinical application.

**Table 1:** A Summary of articles in the literature review that applied deep learning techniques for magnetic resonance imaging involving Crohn’s Disease.

| Author | Journal | Year | Study Design | Database Type | Images evaluated by |
| --- | --- | --- | --- | --- | --- |
| Son et al. | Diagnostic<br>Interventional<br>Radiology | 2023 | Retrospective | Proprietary | Board-certified abdominal<br>radiologists |
| Jeri-McFarlane et al. | Colorectal disease | 2023 | Prospective | Proprietary | N/A |
| Lamash et al. | Journal of<br>Magnetic<br>Resonance<br>Imaging | 2019 | Retrospective | Proprietary | Board-certified radiologist |
| Lian et al. | Results in Physics | 2021 | Retrospective | Proprietary | Board-certified radiologist |
| van Harten et al. | Medical Image<br>Analysis | 2022 | Retrospective | Proprietary | Board-certified radiologist |

Four of the studies were retrospective, one was prospective. In four of the studies, a board-certified radiologist, served as reference standard. No studies have performed external validation.

### Descriptive summary of results

Studies included in this review utilized deep learning techniques for various tasks.

*Son et al*. used deep learning-based reconstruction (DLR) and *Lian et al*. employed CNN to improve image quality. *Lamash et al*. used CNN for inflammation assessment while *Van Harten et al*. measured intestinal motility via centerline segmentation. *McFarlane* et al. utilized 3D image processing and reconstruction (3D-IPR) with AI to create pre-procedure 3D animations of perianal fistulas (**Figure 2**).

*Son et al.* utilized a DLR technique to enhance the quality of MRE images in Crohn’s patients. A qualitative assessment was carried out by two abdominal radiologists who evaluated three distinct sets of images: (1) original, (2) images processed with a conventional filter, and (3) images processed using the deep learning tool.

The mean scores assigned by the radiologists revealed a statistically significant improvement in overall image quality for the DLR image set in comparison to both the original and conventionally filtered images (e.g., Coronal overall image quality: 3.6 (Original), 3.8 (Filtered), 4.7 (DLR)). Additionally, they conducted a signal-to-noise ratio (SNR) analysis and observed a significant increase in SNR when employing the DLR method.

*Lian et. al* developed and tested a CNN algorithm with a dataset from 392 patients with epidemic IBD. For the diagnosis of epidemic IBD, they achieved sensitivity 95% and specificity 47%. This demonstrates their CNN algorithm significantly improved image quality.

*Lamash et al.* employed a 1.5T MRI system and T1-weighted post-contrast VIBE sequence to examine 23 pediatric patients with Crohn’s disease. Their CNN-based segmentation demonstrated Dice Similarity Coefficients of 75% for the lumen, 81% for the wall, and 97% for the background. The median relative contrast enhancement value (P = 0.003) demonstrated discriminatory potential between active and non-active disease segments, while various other extracted markers showed differentiation capacity between segments with and without strictures (P < 0.05).

*Van Harten et. al* utilized a deep learning technique for quantification of intestinal motility as a marker of inflammation. They achieved sensitivity 80%, and PPV 86% for severe bowel disease with 312 annotated segments between the two groups (185 segments in the healthy group vs. 127 segments in the severe bowel disease group).

*McFarlane et al.* evaluated utility of deep learning-based 3D reconstruction models for 4 perineal CD patients. They found the model provided a more comprehensive visual representation of the disease. No quantitative measures were conducted.

### Quality assessment

As per the QUADAS-2 tool, three papers were identified with a high risk of bias in at least one category. Additionally, none of the five papers discussed in this review have been externally validated. A detailed evaluation of the bias risk is provided in **Supplementary Document 2, Table 1 and Table 2**.

**Table 2:**
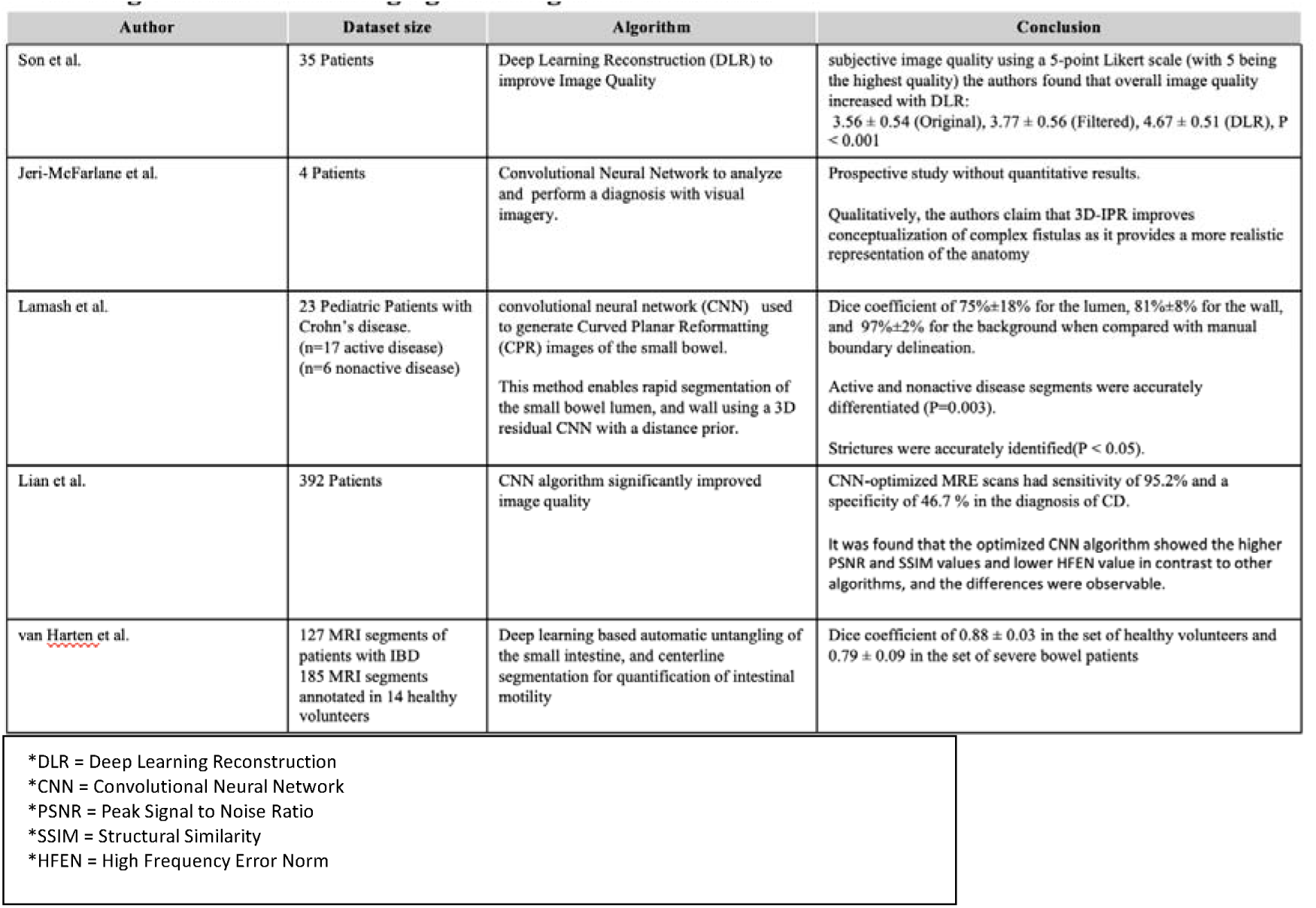
A Summary of articles in the literature review that applied deep learning techniques for magnetic resonance imaging involving Crohn’s Disease.

| Author | Dataset size | Algorithm | Conclusion |
| --- | --- | --- | --- |
| Son et al. | 35 Patients | Deep Learning Reconstruction (DLR) to improve Image Quality | subjective image quality using a 5-point Likert scale (with 5 being the highest quality) the authors found that overall image quality increased with DLR:<br>3.56 ± 0.54 (Original), 3.77 ± 0.56 (Filtered), 4.67 ± 0.51 (DLR), P < 0.001 |
| Jeri-McFarlane et al. | 4 Patients | Convolutional Neural Network to analyze and perform a diagnosis with visual imagery. | Prospective study without quantitative results.<br>Qualitatively, the authors claim that 3D-IPR improves conceptualization of complex fistulas as it provides a more realistic representation of the anatomy |
| Lamash et al. | 23 Pediatric Patients with Crohn's disease.<br>(n=17 active disease)<br>(n=6 nonactive disease) | convolutional neural network (CNN) used to generate Curved Planar Reformatting (CPR) images of the small bowel.<br>This method enables rapid segmentation of the small bowel lumen, and wall using a 3D residual CNN with a distance prior. | Dice coefficient of 75%±18% for the lumen, 81%±8% for the wall, and 97%±2% for the background when compared with manual boundary delineation.<br>Active and nonactive disease segments were accurately differentiated (P=0.003).<br>Strictures were accurately identified(P < 0.05). |
| Lian et al. | 392 Patients | CNN algorithm significantly improved image quality | CNN-optimized MRE scans had sensitivity of 95.2% and a specificity of 46.7 % in the diagnosis of CD.<br>It was found that the optimized CNN algorithm showed the higher PSNR and SSIM values and lower HFEN value in contrast to other algorithms, and the differences were observable. |
| van Harten et al. | 127 MRI segments of patients with IBD<br>185 MRI segments annotated in 14 healthy volunteers | Deep learning based automatic untangling of the small intestine, and centerline segmentation for quantification of intestinal motility | Dice coefficient of 0.88 ± 0.03 in the set of healthy volunteers and 0.79 ± 0.09 in the set of severe bowel patients |

\*DLR = Deep Learning Reconstruction \*CNN = Convolutional Neural Network \*PSNR = Peak Signal to Noise Ratio \*SSIM = Structural Similarity \*HFEN = High Frequency Error Norm

## Discussion

Accurate assessment of CD is pivotal for patient management^19–21^. MRE provides noninvasive insights into both structural and functional aspects without ionizing radiation. Objective endpoints for CD management have been evaluated and established in the form of MRE indices^10,11,22^. However, prior research raises concern about MRE analysis being prone to human error, stemming from the time and attention required in radiologists’ interpretations of the entire bowel ^23^.

The use of deep learning models presents significant potential advancements. It offers enhanced accuracy with these algorithms applied to MRE image analysis for CD^12,13,24^. Dice coefficient values between studies ranged from 0.75 to 0.97, indicating similarity between the results of the deep learning models and those of conventional methods^25,26^.

Recent studies in our review have underscored the capabilities of deep learning in improving MRE evaluation of CD. For example, *Lamash et al*. demonstrated the effectiveness of CNN-based segmentation in pediatric patients, showing high Dice Similarity Coefficients for disease burden characterization and stricture identification ^25^. Their findings indicated that such models may surpass the clinically recommended model in assessing ileal CD activity. This highlights the potential for precise, automated, and non-invasive monitoring of intestinal inflammation in CD patients^27^.

The complex nature of CD can benefit from more reliable assessment of disease activity, distribution, and treatment response. This is especially true for clinical trials, where precise quantification of disease is essential. Interestingly, our review demonstrates varied clinical tasks. These range from improving image quality, segmenting disease to quantify burden, and 3D reconstruction for surgery planning.

Despite the promise shown by these studies, they are not without limitations. All, but one, of the studies are retrospective and none of them have had external validation^24–29^.

Future research needs to include direct comparisons between deep learning and conventional radiological assessments in diverse clinical scenarios. Multicenter prospective studies will be crucial in validating the effectiveness of these AI systems, thereby establishing their role in the clinical management of CD.

In conclusion, deep learning models in CD offer promising enhancements to current MRE readings. Preliminary research indicates acceptable sensitivity and specificity. However, these results are primarily based on retrospective studies and thus require further validation through future research in a clinical setting.

## Data Availability

All data produced in the present work are contained in the manuscript

---

**Supplementary Figure 1:**
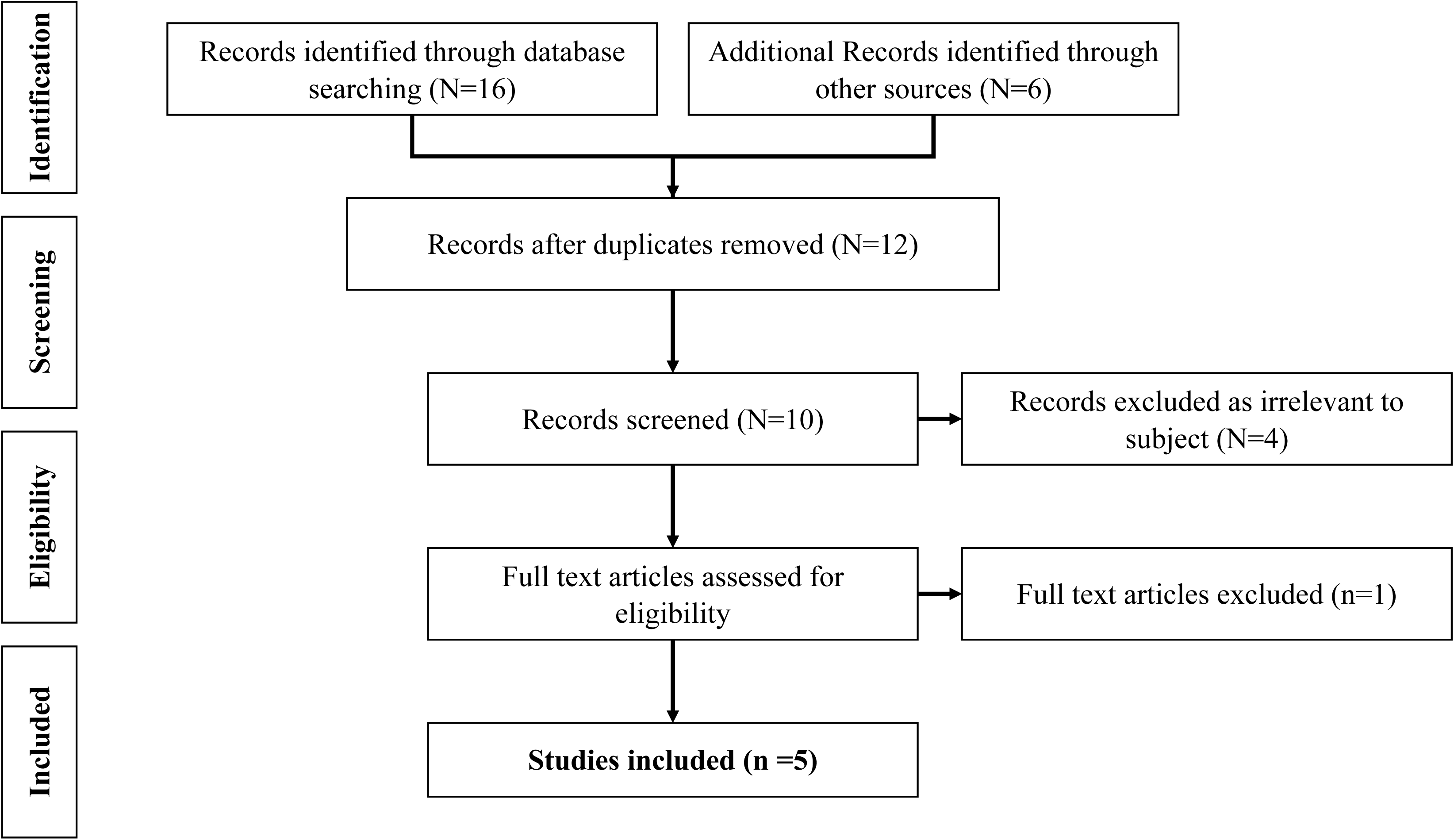
Flow Diagram of the search and inclusion process

