## Supplementary 2 for "Deep Learning in Magnetic Resonance Enterography for Crohn’s Disease Assessment: A Systematic Review"

**Supplementary Online Content**

**Title Page.** Outline………………………………………………………………………….…...1

**Supplementary Material 1.** Literature search strategy…………..……………..…..…..… 2

**Supplementary Table 1**. Quality Assessment of Diagnostic Accuracy Studies-2......…..3

**References**………………………………………………………………………….……….…..4

This supplementary material has been provided by the authors to give readers additional information about the work.

**Supplementary Material 1: Literature search strategy**

Database: Ovid MEDLINE(R) and Epub Ahead of Print, In-Process & Other Non-Indexed Citations and Daily <1946 to December 28, 2023>

Search Strategy:

--------------------------------------------------------------------------------

1 ("mri" OR "MRE" or "magnetic resonance imaging" or "magnetic resonance enterography”)

2 ("crohn's disease" or "crohn's" or "inflammatory bowel disease")

3 ("deep learning" or "convolutional neural networks" or "machine learning" or "artificial intelligence")

***************************

**Supplementary Table 1: Quality Assessment of Diagnostic Accuracy Studies-2 (QUADAS-2) risk of bias assessment**.

Abbreviations: Pt. patient; Ref. reference.
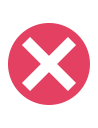
 = high risk of bias;
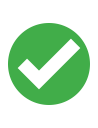
 = low risk of bias.

| **Author** | **Pt. selection^a^** | **Index test^b^** | **Ref. standard^c^** | **Flow and timing^d^** |
| --- | --- | --- | --- | --- |
| Van Harten et al. ^1^ | 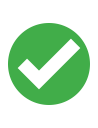 | 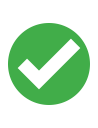 | 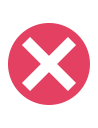 | 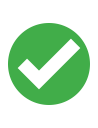 |
| Son et al. ^2^ | 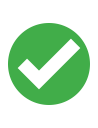 | 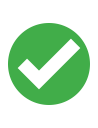 | 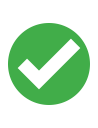 | 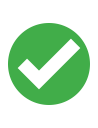 |
| Lamash et al. ^3^ | 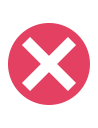 | 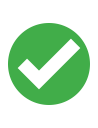 | 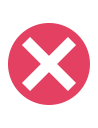 | 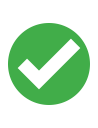 |
| McFarlane et al. ^4^ | 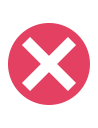 | 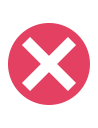 | 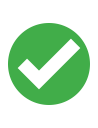 | 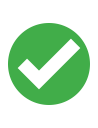 |
| Lian et al. ^5^ | 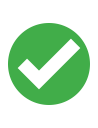 | 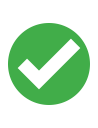 | 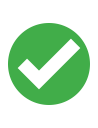 | 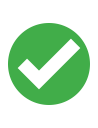 |
